# At-home Testing and Risk Factors for Acquisition of SARS-CoV-2 Infection in a Major US Metropolitan Area

**DOI:** 10.1101/2022.02.02.22269258

**Authors:** Ann E. Woolley, Scott Dryden-Peterson, Andy Kim, Sarah Naz-McLean, Christina Kelly, Hannah H. Laibinis, Josephine Bagnall, Jonathan Livny, Peijun Ma, Marek Orzechowski, Noam Shoresh, Stacey Gabriel, Deborah T. Hung, Lisa A. Cosimi

**Affiliations:** Division of Infectious Diseases, Brigham and Women’s Hospital, Boston, Massachusetts; Department of Immunology and Infectious Diseases, Harvard T.H. Chan School of Public Health, Boston, Massachusetts; Botswana Harvard AIDS Institute, Boston, Massachusetts; Broad Institute of MIT and Harvard, Cambridge, Massachusetts; Department of Molecular Biology and Center for Computational and Integrative Biology, Massachusetts General Hospital, Boston, Massachusetts

**Author notes:** **Corresponding Authors:** Ann Woolley, MD, Division of Infectious Diseases, Brigham and Women’s Hospital, 75 Francis Street, Boston, MA 02115. Work, Lisa Cosimi, MD, Division of Infectious Diseases, Brigham and Women’s Hospital, 75 Francis Street, Boston, MA 02115.

## Abstract

**Importance:** Unbiased assessment of risks associated with acquisition of SARS-CoV-2 is critical to informing mitigation efforts during pandemics.

**Objective:** Understand risk factors for acquiring COVID-19 in a large, prospective cohort of adult residents recruited to be representative of a large US metropolitan area.

**Design:** Fully remote longitudinal cohort study launched in October 2020 and ongoing; Study data reported through June 15, 2021.

**Setting:** Brigham and Women’s Hospital, Boston MA.

**Participants:** Adults within 45 miles of Boston, MA.

**Intervention:** Monthly at-home SARS-CoV-2 viral and antibody testing.

**Main Outcomes:** Between October 2020 and January 2021, we enrolled 10,289 adults reflective of Massachusetts census data. At study entry, 567 (5.5%) participants had evidence of current or prior SARS-CoV-2 infection. This increased to 13.4% by June 15, 2021. Compared to whites, Black non-Hispanic participants had a 2.2 fold greater risk of acquiring COVID-19 (HR 2.19, 95% CI 1.91-2.50; p=<0.001) and Hispanics had a 1.5 fold greater risk (HR 1.52, 95% CI 1.32-1.71; p=<0.016). Individuals aged 18-29, those who worked outside the home, and those living with other adults and children were at an increased risk. Individuals in the second and third lowest disadvantaged neighborhood communities, as measured by the area deprivation index as a marker for socioeconomic status by census block group, were associated with an increased risk in developing COVID-19. Individuals with medical risk factors for severe COVID-19 disease were at a decreased risk of SARS-CoV-2 acquisition.

**Conclusions:** These results demonstrate that race/ethnicity and socioeconomic status are not only risk factors for severity of disease but are also the biggest determinants of acquisition of infection. Importantly, this disparity is significantly underestimated if based on PCR data alone as noted by the discrepancy in serology vs. PCR detection for non-white participants, and points to persistent disparity in access to testing. Meanwhile, medical conditions and advanced age that increase the risk for severity of SARS-CoV-2 disease were associated with a lower risk of acquisition of COVID-19 suggesting the importance of behavior modifications. These findings highlight the need for mitigation programs that overcome challenges of structural racism in current and future pandemics.

**Trial Registration:** N/A

**KEY POINTS:** 

**Question:** What population and occupational groups in the United States are at increased risk for acquiring COVID-19?

**Findings:** In this remote, longitudinal cohort study involving monthly PCR and serology self-testing of 10,289 adult residents of the Boston metropolitan area, 9257 (90.0%) of TestBoston participants acquired evidence of immunity to SARS-CoV-2 through vaccination, infection, or both as of June 15, 2021. Residents identifying as Black, Hispanic/Latinx had an increased risk of acquisition of COVID-19. Healthcare workers were not at increased risk of SARS-CoV-2 acquisition. Individuals with medical risk factors for severe COVID-19 disease were at a decreased risk of SARS-CoV-2 acquisition.

**Meaning:** These results demonstrate that race/ethnicity and socioeconomic status are not only risk factors for severity of disease but also are the biggest determinants of acquisition of infection. These findings highlight the need to address the consequences of structural racism during the development of mitigation programs for current and future pandemics.

## INTRODUCTION

The SARS-CoV-2 pandemic has necessitated frequent decisions regarding prioritization of access to mitigation measures such as testing, contact tracing, housing support, and vaccination among population groups. Considerable disparity in the application of these measures has been observed in the US with decreased uptake among younger adults, racial and ethnic minorities, rural populations, individuals with lower socioeconomic statuses, and certain occupational groups. While several studies have focused on the risk factors for severity of COVID-19 infection, unbiased data are lacking to assess relative population risks regarding the acquisition of COVID-19 which are critical to know to better guide ongoing mitigation efforts by public health authorities, institutions, and health care providers.

While the successful aggregation of SARS-CoV-2 viral polymerase chain reaction (PCR) test results by regional, state, and national public health agencies has permitted comparison of confirmed COVID-19 incidence by geography, age, and racial/ethnic groups, variable access to SARS-CoV-2 testing throughout the pandemic by these same factors, as well as poorer access to testing in many disadvantaged communities, potentially confounds inference related to the risk of different populations acquiring SARS-CoV-2 infection^1,2^. In addition, given that close to half of all COVID-19 infections may be asymptomatic or only mildly symptomatic, thus not prompting PCR testing, public health agency data based on captured viral testing data alone rather than viral and antibody testing may not accurately capture the true incidence. Additionally, the lack of further granularity of testing details such as employment status, household composition, and medical comorbidities, limit the interpretations of local and national testing trends. These confounding factors have likely led to an under reporting of SARS-CoV-2 infections. Consequently, unbiased seroprevalence monitoring is very important for obtaining more accurate estimates of infection and transmission as well as determining the risk factors for acquiring COVID-19. Seroprevalence monitoring also contributes to a more refined estimate of the proportion of individuals who have not yet been exposed to SARS-CoV-2 and are not yet vaccinated, and thus constitute the greatest at-risk group of individuals. Cross-sectional seroprevalence studies have documented that 60-70% of SARS-CoV-2 infections were not clinically detected, but the lack of understanding of the relative timing of infection and potential risks associated with transmission also limits the conclusions of these cross-sectional studies^3,4^.

To inform implementation of risk reduction interventions, we sought to prospectively examine risks associated with the incidence of SARS-CoV-2 infection in a large cohort of adult residents recruited to be representative of the Boston metropolitan area.

## METHODS

By enrolling a large, generalizable cohort of individuals, representative of the metropolitan Boston area, the aim of this study was to understand the risk factors for acquiring COVID-19 and how confirmed cases of COVID-19 and seroprevalence varied across different socioeconomic statuses, racial/ethnic groups, sexes, age groups, medical comorbidities, and occupations between October 2020 and July 2021. In this monthly, cross-sectional study, we tested individuals in the greater metropolitan Boston area for both SARS-CoV-2 PCR as well as SARS-CoV-2 antibodies. In this article, we present results, including seroprevalence estimates and incidence over time, from longitudinally enrolled adult participants between October 2020 to June 15, 2021.

### Study Design

A randomly generated cohort of 150,000 adults 18 years or older who lived within a 45-mile-radius of Boston were eligible to enroll in the TestBoston Study. Participants were offered the opportunity to continue to participate in the longitudinal study with up to 6-months of surveillance testing. The enrolled cohort consisted of individuals normalized to the general population of Massachusetts based on age, gender, and race/ethnicity per state census data^5^.

Baseline surveys were completed by enrolled participants followed by monthly questionnaires to collect information on any new COVID-like symptoms, SARS-CoV-2 testing, and beginning in January 2021, history of COVID vaccination that occurred in the prior 30 days. Self-collection test kits were shipped directly to the participant’s home and included an anterior nasal swab for SARS-CoV-2 PCR and a dried blood spot card and lancet for SARS-CoV-2 IgG serology. Participants shipped the kit back to the Broad Institute in a pre-addressed, pre-paid envelope for processing [Instructions in Appendix]. In addition to monthly surveillance testing irrespective of symptoms, if a study participant developed new symptoms concerning for COVID-19 between scheduled monthly tests or had a known exposure to another individual with COVID-19, the study participant was able to request an additional, ad-hoc test kit.

[Recruitment methods and laboratory details in Appendix.]

### Statistical Analysis

Power analyses set a target enrollment of 10,000 individuals at baseline, followed by 80% of individuals who chose to enroll in the longitudinal portion of the study, assuming a 10% monthly attrition rate during months 3-6. Assuming a baseline seroprevalence of 5%, this sample size was determined to allow us to estimate an overall seroprevalence of COVID–19 infection of 5% (95% CI: 4.5%-5.5%) or higher, and able to estimate the seroprevalence of COVID–19 infection by age groups, gender, and ethnicity with good precision.

Descriptive statistics (proportions and 95% CI, means and standard deviations) were performed for all studied variables: demographics, occupation, socioeconomic status as estimated by area deprivation index^6^, and comorbidities. Additional analyses including multivariate Cox regression analyses were used to explore factors associated with seroprevalence and viral positivity.

## RESULTS

### Participants, testing, and retention

Between October 2020 and January 2021, 102,576 adult residents of metropolitan Boston (estimated population 3,987,800 adults) were invited to join the study and 10,289 (10%) enrolled and completed at least one test kit. The median date of enrollment was November 23, 2020 (interquartile range October 20 to December 23, 2020). 7,181 (70%) of participants were white non-Hispanic, 952 (9.3%) were Hispanic, 925 (9.0%) were Black non-Hispanic, and 889 (8.6%) were Asian non-Hispanic. 5,855 (57%) participants were female. 1765 (17%) were 18 to 29 years old and 1746 (17%) were 65 years old or greater [Table 1]. The TestBoston participants had similar demographic characteristics to the metropolitan Boston population, with some observed differences. The TestBoston cohort included more non-White participants (Black (9% in TestBoston vs 6.7% in metropolitan Boston census data); Hispanic or Latinx (9.3% vs 8.1%); Asian (8.6% vs 6.3%); and multiple and other race/ethnicities (3.3% vs 2.5%)), more females (57% vs 52%), and more participants in professional occupations (healthcare (15% vs 7.2%); education and sciences (12% vs 7%)) and fewer participants in retail, transportation, and other frontline (7.2% vs 26%) than the catchment population. Participants in TestBoston lived in neighborhoods with lower levels of disadvantage and reported being previously diagnosed with COVID-19 at enrollment less frequently than the rate of confirmed COVID-19 in the metropolitan Boston population (2.4% vs 3.5%).

**Table 1.** Baseline Characteristics of TestBoston Participants and Metropolitan Boston Population Abbreviations: ADI, Area Deprivation Index, PCR, polymerase chain reaction Characteristics of TestBoston participants are obtained via self-report and SARS-CoV-2 testing at the time of study entry, with the exception of neighborhood disadvantage which was determined through geolocation of address to census block group (2010 US Census definitions) and Area Deprivation Index (2018 version). Characteristics for metropolitan Boston drawn from resident responses to the 2010 US Census (last available at block group level) in the TestBoston study area (45 mile radius of Boston). Cumulative incidence of COVID-19 for towns and cities within the TestBoston study area obtained from public reporting by the Massachusetts Department of Public Health.

| Characteristic | TestBoston, N = 10,289 | Metro Boston, N = 3,987,800 |
| --- | --- | --- |
| **Sex** |  |  |
| *Female* | 5,855 (57%) | 2,090,300 (52%) |
| *Male* | 4,434 (43%) | 1,897,500 (48%) |
| **Age** |  |  |
| *18 to 29 years* | 1,765 (17%) | 890,600 (22%) |
| *30 to 44 years* | 3,244 (32%) | 1,036,200 (26%) |
| *45 to 64 years* | 3,534 (34%) | 1,394,400 (35%) |
| *65 years and older* | 1,746 (17%) | 666,600 (17%) |
| **Race and ethnicity** |  |  |
| *White* | 7,181 (70%) | 3,045,800 (76%) |
| *Black* | 925 (9.0%) | 267,900 (6.7%) |
| *Hispanic or Latinx* | 952 (9.3%) | 324,100 (8.1%) |
| *Asian* | 889 (8.6%) | 250,400 (6.3%) |
| *Multiple and other races/ethnicities* | 342 (3.3%) | 99,600 (2.5%) |
| **Neighborhood disadvantage (ADI quintiles)** |  |  |
| *0 to 20 percentile (lowest disadvantage)* | 4,320 (42%) | 965,400 (24%) |
| *20 to 40 percentile* | 3,052 (30%) | 983,000 (25%) |
| *40 to 60 percentile* | 1,896 (18%) | 888,900 (22%) |
| *60 to 80 percentile* | 799 (7.8%) | 729,600 (18%) |
| *80 to 100 percentile (highest disadvantage)* | 222 (2.2%) | 420,900 (11%) |
| **Occupation** |  |  |
| *Business, legal, managerial* | 3,252 (32%) | 1,112,700 (28%) |
| *Healthcare* | 1,526 (15%) | 285,900 (7.2%) |
| *Retail, transportation, and other frontline* | 745 (7.2%) | 1,037,500 (26%) |
| *Education and sciences* | 1,199 (12%) | 277,300 (7.0%) |
| *Arts, sports, and other* | 742 (7.2%) | 170,400 (4.3%) |
| *Retired, unemployed, or student* | 2,825 (27%) | 1,103,800 (28%) |
| **Household members** |  |  |
| *Living alone* | 1,314 (13%) | --- |
| *Living with other adults* | 5,645 (55%) | --- |
| *Living with children* | 3,330 (32%) | --- |
| **Location of work** |  |  |
| *Home or remote* | 7,185 (70%) | --- |
| *In-person* | 3,104 (30%) | --- |
| **Medical conditions** |  |  |
| *No major comorbidities* | 5,387 (52%) | --- |
| *Immunocompromise* | 862 (8.4%) | --- |
| *Cardiovascular, pulmonary disease, or diabetes* | 3,434 (33%) | --- |
| *Psychiatric disorder* | 606 (5.9%) | --- |
| **Prior COVID-19** |  |  |
| *No Prior COVID-19* | 9,717 (94%) | 5,274,300 (97%) |
| *PCR confirmed COVID-19* | 245 (2.4%) | 189,800 (3.5%) |
| *Serology-detected COVID-19* | 327 (3.2%) | --- |

Through the analysis date of June 15, 2021, TestBoston participants were followed for 5174 person-years (median 6.2 months). Retention for 4 months or more was 56%. During this period, a total of 44,886 viral PCR and 43,628 serology tests were performed by TestBoston and 15,090 viral PCR tests performed outside of TestBoston were reported by participants. Of the TestBoston tests performed between October 2020 and June 2021, 252 (0.6%) were positive PCR tests and 12,633 (29%) were positive serology tests.

### SARS-CoV-2 infection and vaccination

At the time of study entry, 459 (4.9%) participants had serologic evidence of prior SARS-CoV-2 infection, 2.7 times higher than the 173 who reported having known prior infection. An additional 66 participants were diagnosed by positive PCR at entry. During longitudinal follow-up, an additional 719 (6.9%) infections were detected with 126 by PCR testing outside of TestBoston, 157 by TestBoston PCR testing, and 613 by serologic testing. The detection of SARS-CoV-2 among TestBoston closely followed the epidemic curve for COVID-19 in metropolitan Boston [Appendix Figure] with peak incidence in January 2021.

The cumulative incidence of SARS-CoV-2 infection among TestBoston participants based on serologic testing increased from 5.1% on December 1, 2020 to 12.7% on June 15, 2021. Standardized by age, race/ethnicity, and neighborhood disadvantage, the estimated cumulative incidence among adults in metropolitan Boston increased from 5.1% (95% CI 4.3 to 5.9%) in December 2020 to 12.8% (95% CI 11.9 to 13.7%) in June 2021. In comparison, the cumulative incidence of confirmed COVID-19 in metropolitan Boston, as reported by the Massachusetts Department of Public Health, was 3.1% in December 2020 and 8.4% in June 2021. Non-White TestBoston participants were more likely to have SARS-CoV-2 infection only identified via serology (ie, not clinically recognized or recorded).

SARS-CoV-2 vaccination became available to some participants beginning in December 2020 and with increasing eligibility through early 2021. A total of 8879 (86.3%) participants received at least one vaccine dose during the study period. By June 2021, 9257 (90.0%) of TestBoston participants acquired evidence of immunity to SARS-CoV-2 through vaccination, infection, or both [Figure 2]. Standardized to the population, an estimated 89% (95% CI 88 to 91%) of adult residents acquired evidence of immunity to SARS-CoV-2.

**Figure 1.**
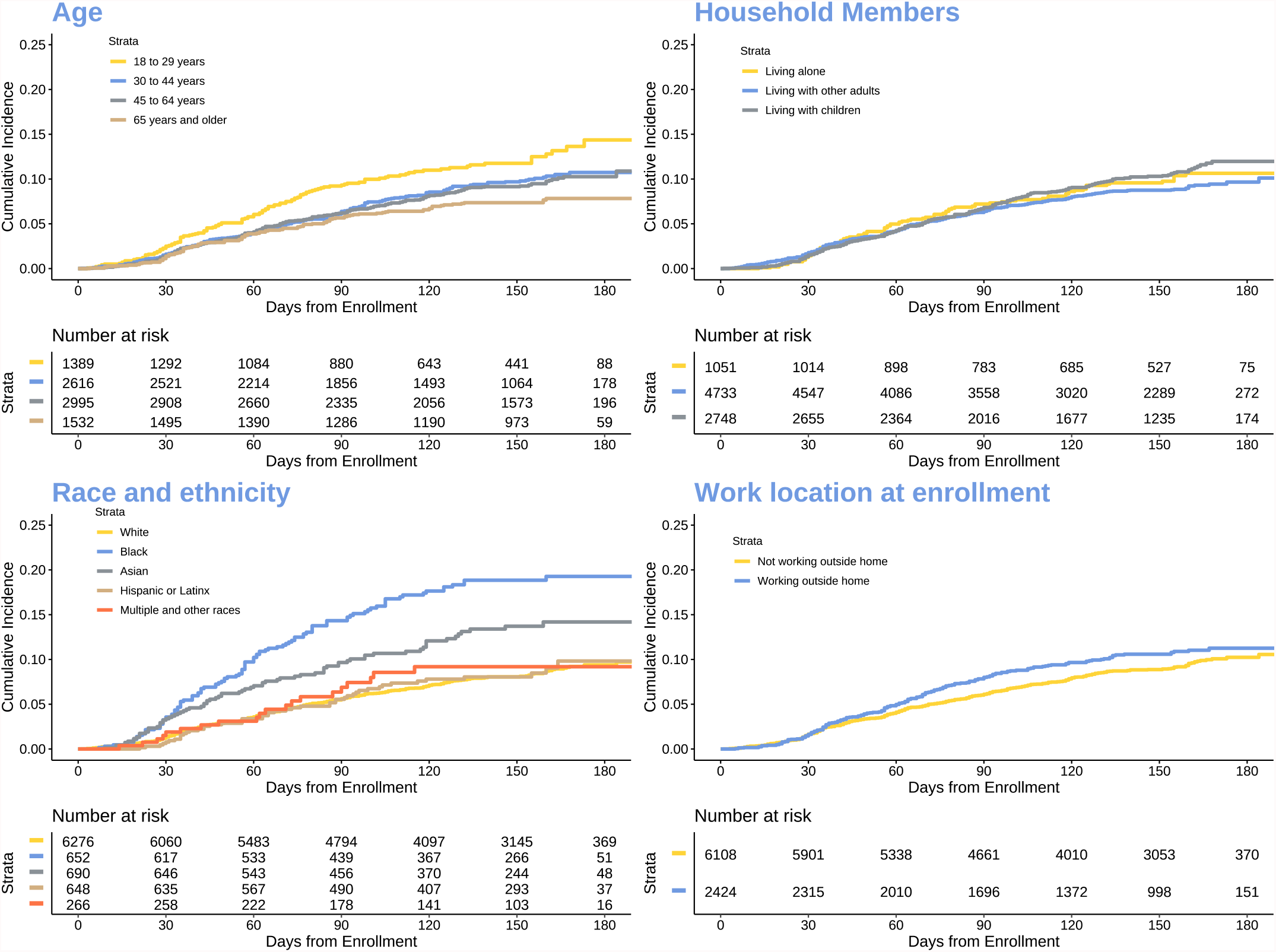
Comparison of Cumulative Incidence of SARS-CoV-2 Infection Among TestBoston Participants Without Prior Infection at Study Entry Note: Characteristics as reported by participants at study entry.

**Figure 2.**
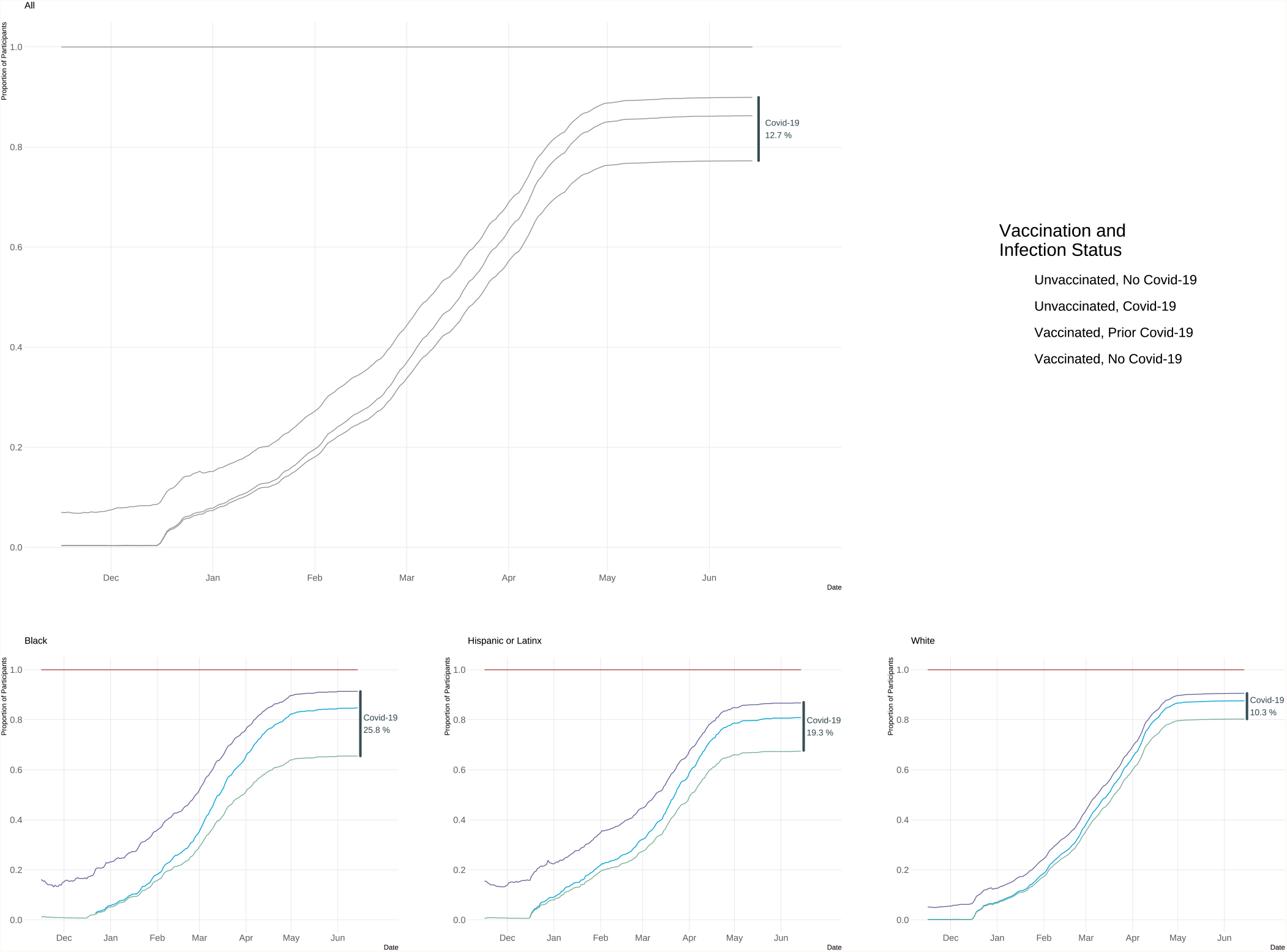
Cumulative Infection and Vaccination Among TestBoston Participants Note: Date of first positive COVID-19 test (PCR or serology) and first dose of a SARS-CoV-2 vaccine. Participant-reported race and ethnicity at study entry. ⍰

### Risk of Incident SARS-CoV-2 Infection

Among 8532 TestBoston participants without evidence of prior SARS-CoV-2 infection at study entry, 717 (8.4%) acquired COVID-19 during the study period. A multivariate Cox regression analysis [Table 2] showed individuals of ages 30-44 (HR 0.76, 95% CI 0.66-0.86; p=<0.001), 45-64 (HR 0.72, 95% CI 0.63-0.81; p=<0.001), and 65 years and older (HR 0.72, 95% CI 0.61-0.85; p=<0.001) had a significantly lower incidence of SARS-CoV-2 infection compared to 18– 29-year-olds. Conversely, Black non-Hispanic, Asian non-Hispanic, and Hispanic participants had a significantly higher incidence. Black non-Hispanics had a 2.2 greater risk compared to white non-Hispanics (HR 2.19, 95% CI 1.91-2.50; p=<0.001). Hispanics had a 1.5 greater risk (HR 1.52, 95% CI 1.32-1.71; p=<0.016) and Asian non-Hispanics had a 1.22 greater risk (HR 1.22, 95% CI 1.04-1.43; p=0.001) compared to white non-Hispanics. Men had a 16% risk reduction in developing COVID-19 relative to woman (HR 0.84, 95% CI 0.77-0.92; p=<0.001).

**Table 2.** Multivariable Stratified Cox Proportional Hazards Regression Analysis for Incidence of SARS-CoV-2 Infection

| Characteristic | HR<sup>1</sup> | 95% CI<sup>1</sup> | p-value |
| --- | --- | --- | --- |
| **Sex** |  |  |  |
| Female | Ref. | Ref. |  |
| Male | 0.84 | 0.77, 0.92 | **<0.001** |
| **Age** |  |  |  |
| 18 to 29 years | Ref. | Ref. |  |
| 30 to 44 years | 0.76 | 0.66, 0.86 | **<0.001** |
| 45 to 64 years | 0.72 | 0.63, 0.81 | **<0.001** |
| 65 years and older | 0.72 | 0.61, 0.85 | **<0.001** |
| **Race and ethnicity** |  |  |  |
| White | Ref. | Ref. |  |
| Black | 2.19 | 1.91, 2.50 | **<0.001** |
| Hispanic or Latinx | 1.52 | 1.32, 1.75 | **<0.001** |
| Asian | 1.22 | 1.04, 1.43 | **0.016** |
| Multiple and other races/ethnicities | 1.06 | 0.82, 1.37 | 0.66 |
| **Neighborhood disadvantage (ADI quintiles)** |  |  |  |
| 0 to 20 percentile (lowest disadvantage) | Ref. | Ref. |  |
| 20 to 40 percentile | 1.01 | 0.91, 1.12 | 0.86 |
| 40 to 60 percentile | 1.17 | 1.04, 1.31 | **0.007** |
| 60 to 80 percentile | 1.38 | 1.19, 1.60 | **<0.001** |
| 80 to 100 percentile (highest disadvantage) | 0.89 | 0.66, 1.20 | 0.44 |
| **Medical conditions** |  |  |  |
| No major comorbidities | Ref. | Ref. |  |
| Immunocompromise | 0.97 | 0.83, 1.13 | 0.70 |
| Cardiovascular, pulmonary disease, or diabetes | 0.91 | 0.83, 1.00 | **0.049** |
| Psychiatric disorder | 0.71 | 0.58, 0.88 | **0.002** |
| **Location of work** |  |  |  |
| Home or remote | Ref. | Ref. |  |
| In-person | 1.32 | 1.19, 1.47 | **<0.001** |
| **Occupation** |  |  |  |
| Business, legal, managerial | Ref. | Ref. |  |
| Healthcare | 1.04 | 0.90, 1.21 | 0.58 |
| Retail, transportation, and other frontline | 1.02 | 0.86, 1.20 | 0.83 |
| Education and sciences | 0.69 | 0.59, 0.80 | **<0.001** |
| Arts, sports, and other | 1.08 | 0.93, 1.27 | 0.30 |
| Unemployed | 1.06 | 0.94, 1.19 | 0.35 |
| **Household members** |  |  |  |
| Living alone | Ref. | Ref. |  |
| Living with other adults | 0.92 | 0.81, 1.04 | 0.19 |
| Living with children | 1.19 | 1.04, 1.37 | **0.013** |
| **Vaccination** |  |  |  |
| Unvaccinated | Ref. | Ref. |  |
| Vaccinated (0 to 30 days) | 0.08 | 0.05, 0.11 | **<0.001** |
| Vaccinated (31 or more days) | 0.01 | 0.00, 0.02 | **<0.001** |

<sup>1</sup>HR = Hazard Ratio, CI = Confidence Interval
Abbreviations: ADI, Area Deprivation Index; HR, hazard ratio; CI, confidence interval.
Model stratified by month of study entry to limit temporal confounding in the context of varying epidemic intensity during enrollment period. Vaccination status is time-updated and other characteristics modeled as fixed as measured at time of study entry.

Individuals in the second and third lowest disadvantaged neighborhood communities, as measured by the area deprivation index (ADI) as a marker for socioeconomic status by census block group, were associated with an increased risk in developing COVID-19 (HR 1.17, 95% CI 1.04-1.31; p=0.007 and HR 1.38, 95% CI 1.19-1.60; p=<0.001). Living alone was associated with a lower risk of getting COVID-19 as households with adults and children had an increased risk in developing COVID-19 compared to those who lived alone (HR 1.19, 95% CI 1.04-1.37; p=0.013). Individuals who work outside of the home had an increased risk of acquiring COVID-19 compared to those who worked remotely (HR 1.32, 95% CI 1.19-1.47; p=<0.001). Those who worked in education had a lower adjusted risk of developing COVID-19 (HR 0.69, 95% CI 0.59-0.80; p=<0.001). Frontline workers did not have an adjusted increased risk (HR 1.02, 95% CI 0.86-1.20; p=0.83). Vaccination showed a 97.2% effectiveness during this study period (HR 0.01, 95% CI 0.00-0.02; p=<0.001).

In contrast to medical risk factors for increased severity of infection, individuals with major medical comorbidities did not have an increased risk of acquiring SARS-CoV-2. In fact, individuals with cardiovascular disease, pulmonary disease, obesity, or diabetes had a lower adjusted risk of acquiring COVID-19 (HR 0.91, 95% CI 0.83-1.00; p=0.049) as did individuals with underlying psychiatric disorders such as depression and anxiety (HR 0.71, 95% CI 0.58-0.88; p=0.002).

## DISCUSSION

TestBoston is one of the largest longitudinal, remote, at-home direct-to-patient COVID-19 testing studies in the United States, established to follow a cohort of over 10,000 individuals who were representative of the residents of Boston, a US metropolitan city with a diverse population, in order to assess the risk factors associated with the development of SARS-CoV-2 infection. There are significant impediments to participating in research studies that disproportionately impact the most vulnerable individuals, particularly those of Black race and Hispanic/Latinx ethnicity, due to language barriers, mistrust in medical research, and time and travel commitments that cannot be accommodated by shift workers, in particular, and those of lowest socioeconomic status. Therefore, this study was designed as a fully remote study that was equitable, scalable, and allowed for a wide geographic catchment area that enabled recruitment and retention of those individuals who are often not represented in these research studies.

As of June 15, 2021, 90% of individuals living in the greater Boston area had either previously been infected by SARS-CoV-2 and/or were vaccinated. While reinfection can occur despite prior infection and vaccine breakthrough infections occur, those 90% of individuals are much less likely to be at risk of severe COVID-19 were they to now acquire SARS-CoV-2 infection^7–11^. Among the remaining individuals who are therefore at higher risk for severe COVID-19 infection, Black and/or Hispanic individuals are disproportionately represented. This disparity is due to considerably lower vaccination rates in Black and Hispanic individuals, as their rates of prior COVID-19 infection were higher than those of other racial and ethnic groups.

Black race, Asian race, and Hispanic ethnicity were strongly associated with seropositivity and COVID-19 cases, echoing broader racial inequities in testing and consistent with previous reports on the disproportionate burden of disease among some racial and ethnic subgroups seen in the US during the COVID-19 pandemic^12–14^. Here, we now confirm that this disparity is significantly underestimated if based on SARS-CoV-2 PCR testing data alone, likely due to unequal access to testing. While there was a 2-fold greater positivity rate based on serology compared to PCR testing for white non-Hispanic and Hispanic individuals, there was a 3.5-fold and 4-fold difference for Black and Asian non-Hispanic individuals.

Twenty-six percent of Black non-Hispanics were seropositive for SARS-CoV-2 prior to vaccination, but only 7% had tested positive at any time via viral PCR testing either during the study or prior to study enrollment. Conversely, only 9% of white non-Hispanic individuals were SARS-CoV-2 seropositive prior to vaccination and nearly half of those cases had been detected via viral PCR testing suggesting greater access to testing for white non-Hispanic individuals in the greater Boston area. This disparity in testing is consistent with what has been seen throughout other metropolitan cities throughout the US, showing the marginalization of majority Black areas due to economic disenfranchisement and limited health-promoting attributes^1,15–17^. Notably in New York City, efforts to increase testing availability to decrease the disparity in the placement of testing sites has begun to make an impact^17^. It is imperative that access to testing is improved for vulnerable groups and that racial and ethnic disparities in access to testing are decreased. This is critical for identifying cases and preventing the spread of COVID-19 among these groups. TestBoston has demonstrated that at-home testing is one way to decrease barriers and expand testing^18^.

Disparities in testing result in disparities in recognizing cases of COVID-19. The early identification of cases when an individual is asymptomatic or only mildly symptomatic is critical in preventing severe COVID-19 infection and associated morbidities and mortality now that treatment options such as monoclonal antibodies are available to decrease an individual’s risk of severe COVID-19 and hospitalization. These study results highlight the importance of understanding the risk factors associated with the acquisition of SARS-CoV-2 infection and the associated negative impact that testing inequities has on individuals and certain communities receiving the needed medical interventions to prevent ongoing and more severe disease.

We know in theory that an individual’s medical comorbidities and behaviors can affect the risk of acquisition of COVID-19, but the data to date has focused more on the risk factors for severity of disease rather than on the risk factors for acquisition. Risk factors of acquisition are critical to elucidating who we should target for mitigation efforts as well as who we should focus the emerging outpatient treatment interventions, including monoclonal antibodies as well as oral antivirals as they become available, to attenuate the severity of disease, ongoing transmission to those individuals who are at higher risk for hospitalization, and minimize long-covid^19^.

Younger aged individuals (18-29 years old) had a higher adjusted risk of acquiring COVID-19 compared to individuals who were 30 years old or older which may have reflected social behaviors during this period of the pandemic. However, the older age groups had a lower proportion of COVID-19 cases detected by PCR testing and a greater proportion of cases detected only by antibody testing prior to vaccination. This highlights the greater access to viral testing programs that were present at colleges and universities, and certain companies or places of work. This has direct negative clinical impacts as those who are older and have a higher risk of severity of disease were less likely to have had viral COVID-19 testing which significantly disadvantages them in terms of potential outpatient treatment options to prevent hospitalizations.

Surprisingly, while cardiovascular and pulmonary comorbidities including obesity, diabetes, asthma, and chronic obstructive pulmonary disease as well as immunosuppressive conditions such as hematologic malignancy, solid organ transplant, or being on immunosuppressive medications, increase an individual’s risk for severe illness from SARS-CoV-2 infection, this study found that these medical conditions do not increase one’s risk of acquiring COVID-19. Having a psychiatric disorder was seen to be associated with lower risk of acquiring COVID-19, in contrast to severe disease, suggesting that the risk of acquisition can be mitigated by behavior. Strengths of this study include the frequent, repeat paired SARS-CoV-2 viral and antibody testing of a wide range of individuals from differing racial and ethnic groups, ages, socio-economic statuses, occupations, and household compositions that are representative of a greater metropolitan city, Boston. This granularity of detail on such a large representative and unbiased cohort sets this study apart and expounds upon the more limited data that are monitored at the city, state, and national levels. Our study design avoided the biases that have been seen in many of the other large COVID-19 studies because it was not limited to participants who were only testing due to being symptomatic, having risk factors for developing severe covid, or presenting with severe disease. We were mindful that data quality matters as much, if not more, than data quantity and ensured that our findings were not a victim of the “big data paradox,” a mathematical tendency of big data sets to minimize errors due to small sample size while magnifying errors linked to systematic biases that make the sample a poor representation of the larger population^20,21^. As a result of our unbiased study design that captured the demographics and composition of a metropolitan US city, we were able to learn key findings about the risks of acquisition of SARS-CoV-2 infection at an individual as well as a community level. Limitations of this study include the fact that the results analyzed for this study stop in mid-June 2021, prior to the Delta and now Omicron variants having become the predominant circulating strains in this geographic area. Other limitations include that asymptomatic viral PCR testing was only offered once monthly and the inability to detect new cases via serology testing post-vaccination.

The COVID-19 pandemic has further highlighted the social and racial inequities that exist in health care and has brought these discussions to the forefront of public health. It has highlighted that health equity is still not a reality as COVID-19 has unequally affected many racial and ethnic minority groups. Testing only captured a fraction of the cases which resulted in an immense inequity of counting of cases in certain populations. As we face SARS-CoV-2 becoming an endemic viral infection, it is paramount to continue understanding the risk factors for acquisition rather than just severity of illness and which of those factors can be mitigated as preventing acquisition of infection is the best intervention to prevent severe illness. These data also demonstrate that a major determinant of risk of acquisition is behavior and that a major risk-reducing intervention is behavioral modification. For example, in younger individuals, behavior dominates their risk for acquisition, while in individuals with significant medical comorbidities, any possibility of increased acquisition may be offset by active behavioral modifications to minimize exposure. Thus, the importance of behavioral modification during a pandemic cannot be overemphasized, in contrast to medical comorbidities and older age which cannot be modified.

Taken together, these results suggest both that the consequences of structural racism on risk must be considered during the development of mitigation programs for current and future pandemics and that an important component of these mitigation programs must be to educate and encourage risk modifying behaviors. While the actual risk factors may change as the status of the pandemic continues to evolve, including waning immunity from vaccination, the rise of variants, the availability of testing, and the changing behavioral norm of local communities and nations, these two principles will likely remain firm for the foreseeable future.

## Data Availability

All data produced in the present study are available upon reasonable request to the authors.

## Acknowledgements

The work was supported by Brigham and Women’s Hospital COVID Response Fund, the Klarman Family Foundation, Skoll Foundation, ThermoFisher, the Helgeson Family Foundation, additional anonymous funders, as well as the Harvard University Center for AIDS Research (CFAR), a funded program of the National Institutes of Health (P30 AI060354). The contents of this manuscript are solely the responsibility of the authors and do not necessarily represent the official views of the National Institutes of Health or the institutions with which the authors are affiliated. The funding source had no role in the design and conduct of the study; collection, management, analysis, and interpretation of the data; preparation, review, or approval of the manuscript; and decision to submit the manuscript for publication. Drs. Woolley, Dryden-Peterson, and Cosimi had full access to all the data in the study and take responsibility for the integrity of the data and the accuracy of the data analysis. We would like to thank Dr. Lawrence C. Madoff and Catherine Brown at the Massachusetts Department of Public Health for their support of this study and valuable input.

## Notes

### Competing Interest Statement

The authors have declared no competing interest.

### Funding Statement

This study was funded by Brigham and Womens Hospital COVID Response Fund, the Klarman Family Foundation, Skoll Foundation, ThermoFisher, the Helgeson Family Foundation, additional anonymous funders, as well as the Harvard University Center for AIDS Research (CFAR), a funded program of the National Institutes of Health (P30 AI060354).

### Author Declarations

IRB of Mass General Brigham gave ethical approval for this work.

