## Supplemental Appendix for "At-home Testing and Risk Factors for Acquisition of SARS-CoV-2 Infection in a Major US Metropolitan Area"

Instructions:

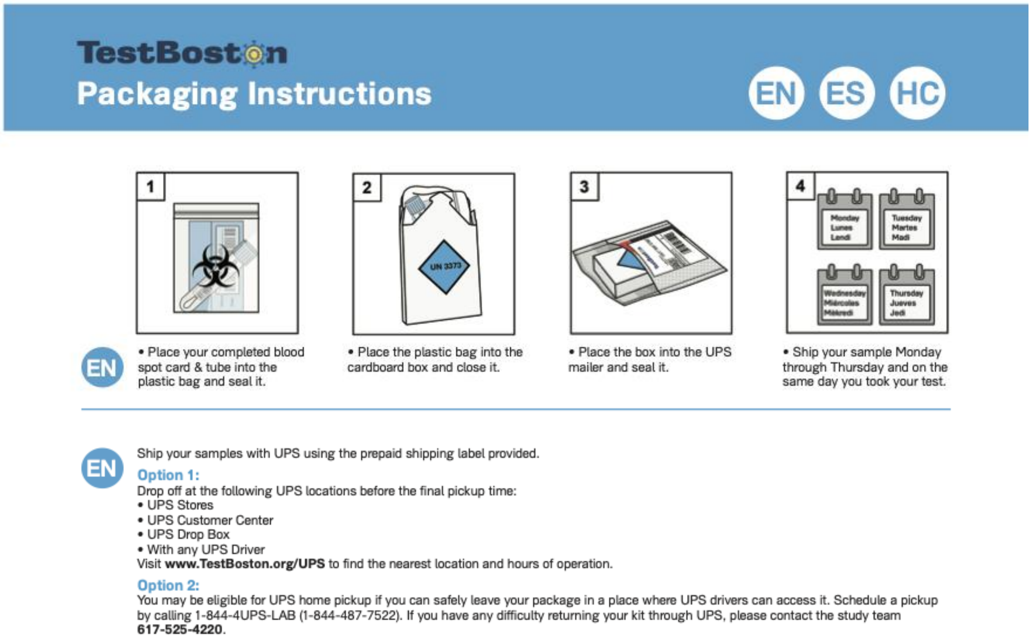

TestBoston

617-525-4220

www.TestBoston.org

EN **Important:** Before collecting your samples, please schedule your pickup or locate your nearest dropbox. To ensure the accuracy of your test results, it is important to mail your sample on the **same day it is collected.**

### TestBoston
#### Nasal Swab Kit Directions

EN

**IMPORTANT:**
**Your specimen must be shipped on a Monday, Tuesday, Wednesday or Thursday:**
- **Ship your specimen on the same day you take your test.**
- **Do not collect your specimen until you confirm the final pickup time of your UPS dropbox or have scheduled a home pick-up.**
- **Shipping instructions can be found on the back of the booklet and at www.testboston.org/ups**

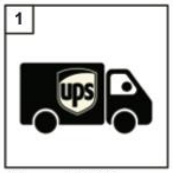

• Plan your UPS pickup or drop off

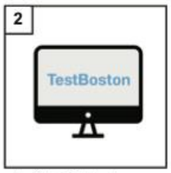

• Log into **TestBoston.org** to complete your Symptom Survey when you are ready to take your test.

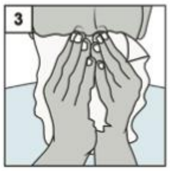

• Blow your nose.

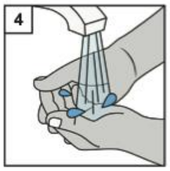

• Wash and dry your hands.

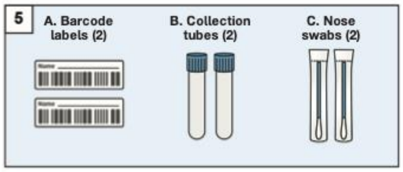

• Unpack your kit and place all the contents on a clean, dry surface.

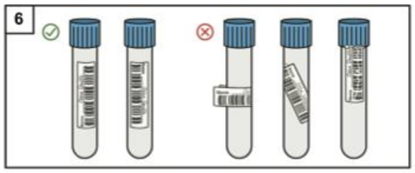

• On two labels (A), write your name and stick a label on the side of each collection tube (B), as shown.

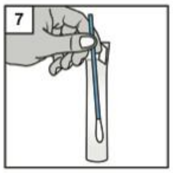

• Take the swab (C) out of its package.
• Do not touch the tip of the swab.

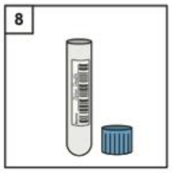

• Pull off the top of the collection tube (B).

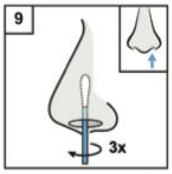

• Rotate the swab in a circle around the inside edge of your nostril at least 3 times and hold in place for 15 seconds.

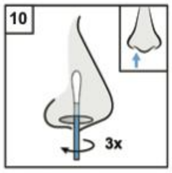

• Move the swab to your other nostril and repeat step 9.

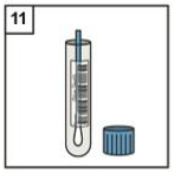

• Put the swab into the collection tube so that the soft tip is at the bottom of the tube.

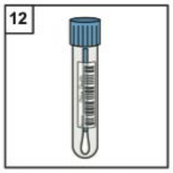

• Push on the top of the collection tube.
• Repeat steps 7-12 with the second tube.
• Wash and dry your hands.

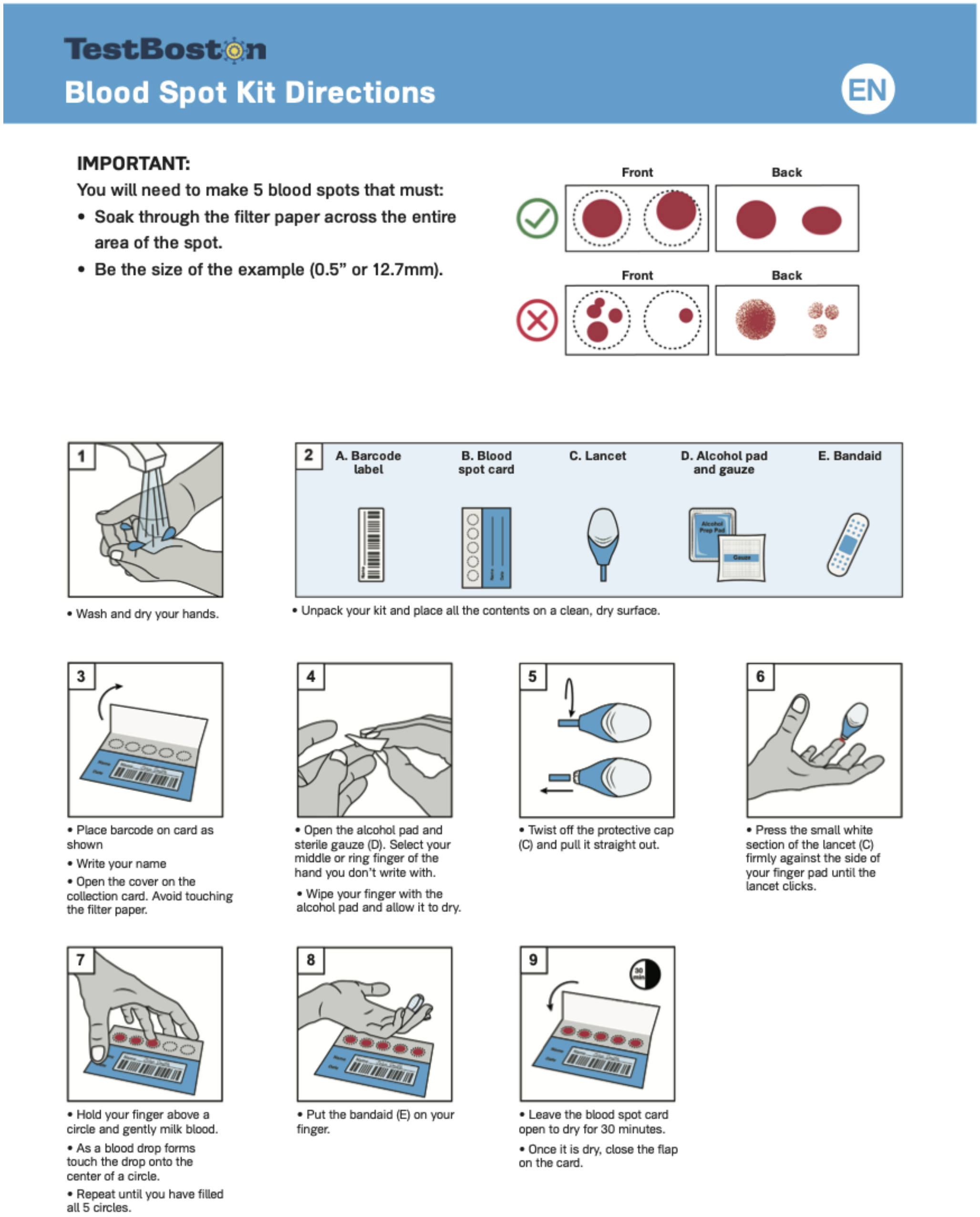


Recruitment methods:

A randomly generated cohort of 150,000 adults 18 years or older who received medical care at Massachusetts General Brigham (MGB), a not-for-profit, integrated healthcare system with 14 affiliated hospitals, and live within a 45-mile-radius of Boston, were eligible to enroll in the TestBoston Study. In addition, individuals who had not received medical care at MGB but learned about the study elsewhere and expressed interest in participating were also eligible.

Between October 1, 2020 and February 2021, 102,576 people were invited to join the study, of which, 10,289 (10.0%) enrolled and returned at least one of six possible test kit. "Direct invitations" were sent via mail and/or electronically to 12,758 individuals who previously opted-in to be contacted about MGB research studies of whom 1,848 (14%) enrolled. "Physician invitations" were sent to 85,505 individuals of whom 5584 (6.5%) enrolled. "Volunteer invitations" were sent to 4,313 individuals (with or without prior connection to MGB), of which 2752 (64%) enrolled.

Laboratory Details:

*Anterior Nasal swabs for SARS-CoV-2 PCR*

Self-collected anterior nasal swab samples were returned via overnight shipping and processed at the Broad Institute's CLIA certified Clinical Research Sequencing Platform (CRSP) lab which has Emergency Use Authorization approval from the Federal Drug Administration to perform high-throughput PCR testing of dry anterior nasal swabs for SARS-CoV-2. Viral swabs were processed in parallel with clinical samples received from local hospitals and public health departments. All results were reported automatically to the Massachusetts Department of Public Health through the Massachusetts Virtual Epidemiologic Network (MAVEN).

*Dried Blood Spot for SARS-CoV-2 IgG Spike Receptor Binding Domain*

Whatman cards containing self-collected dried blood spots labeled with individualized bar codes were run in the Broad Institute's Microbial 'Omics Core (MOC) for high-throughput, partially automated enzyme-linked immunosorbent assay (ELISA) targeting SARS-CoV-2 antigen(s) to identify serological evidence of IgM and IgG antibodies. Results of serology testing were reported in aggregate to the study participants on the publicly available TestBoston data dashboard as well as to key stakeholders including the Massachusetts Department of Public Health.

*Test Boston ELISA method*

Quantitative ELISAs to measure antibodies to the receptor binding domain (RBD) and nucleocapsid (N) proteins (BROAD RBD and BROAD NC, respectively) were developed and performed at the Broad Institute (Cambridge, MA). From each DBS sample, a 3-mm dry blood spot (DBS) was punched into a well in a Slicprep 96 Device (Promega, cat# V1391). 440 µL dilute buffer was added to each well and incubate at 4 °C overnight with shaking. The second day morning, 50 µL DBS eluate were added to MaxiSorp 384-well microplates (Sigma) pre-coated with 50 µL/well of 2,500 ng/ml of SARS-CoV-2 RBD or 50 µL/well of 1,000 ng/ml of SARS-CoV-2 N protein, and incubated for 30 min at 37 °C. Plates were then washed and 50 µL/well of 1:25,000 diluted detection antibody solution (HRP-anti human IgG, Bethyl Laboratory) was added. After an incubation for 30 min at RT, plates were washed and 40 µL/well of Pierce TMB peroxidase substrate (ThermoFisher) was added. The reaction was stopped by adding 40 µL/well of stop solution (0.5 M H2SO4). The OD was read at 450 nm and 570 nm on a BioTek Synergy HT. Antibody concentrations were estimated and normalized using standard curves generated from duplicate serial dilutions of CR3022 IgG1 (Absolute Antibody) for RBD and an IgG positive plasma serum sample for N.

Statistical Analysis:

*Additional Details to the Methods Section in the Manuscript*

Comparing percent SARS-CoV-2 IgG positivity by racial/ethnic group, the sample size allowed us to detect an odds ratio of 2.0 or more at baseline, with 84.5-100% power if the prevalence was 5% or higher and with 82.5-99.9% at the 6-month visit if the prevalence was 10% or higher. Based on simulations using a simple susceptible-exposed-infected-recovered model for viral spread, we chose a population size such that if 10% of our longitudinal cohort was IgG positive at the outset, assuming an R0 = 1.5 over our study period, which presumed partial effectiveness of social distancing and other community interventions over this period, we would have >95% power to detect a 2-fold difference in susceptibility. This left some margin for detecting a difference even if viral spread through the population was very slow: if R0 = 1.1, we would still have 80% power to detect a 10-fold difference in susceptibility, and 50% power to detect a 3-fold difference in susceptibility.

Figure:

Daily Positive COVID-19 PCR Tests of TestBoston Participants (Blue Bars) and Daily Incidence in Greater Boston Area (Green Line)

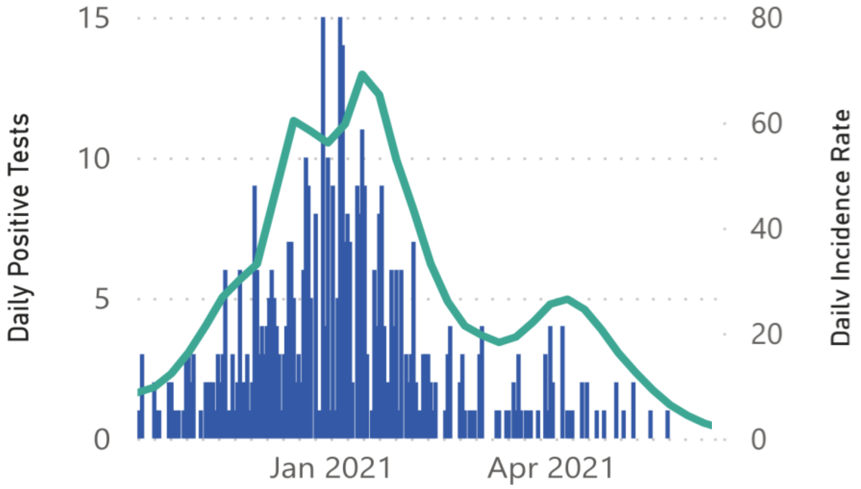
